# Impact of armed conflict on measles surveillance and zero-dose vaccination status in Tigray, Ethiopia, 2018 - 2024

**DOI:** 10.64898/2026.01.29.26345160

**Authors:** Habtamu Milkias Wolde, Youngoh Bae, Ali Raza, Seung Won Lee

## Abstract

**Background:** Armed conflict disrupts health systems, undermining routine immunization and disease surveillance. These disruptions can delay outbreak detection and allow population susceptibility to accumulate unnoticed. This study assessed the impact of the 2020 - 2022 conflict in Tigray, Ethiopia, on measles epidemiology, focusing on surveillance reporting, age distribution of cases, and vaccination status.

**Methods:** We conducted a retrospective longitudinal analysis of national case-based measles surveillance data from Ethiopia (2018–2024; n = 69,866). The study period was classified into pre-conflict (2018 - 2019), conflict peak (2020 - 2022), and post-conflict recovery (2023 - 2024) phases. Two-way analysis of variance examined regional differences in age at infection across phases. Multivariable logistic regression estimated adjusted odds ratios (aORs) for confirmed measles cases being unvaccinated (zero-dose), using the pre-conflict period as the reference and adjusting for age and sex. Surveillance quality was assessed using demographic data completeness.

**Results:** During the conflict peak, reported measles cases from Tigray declined to 0.01% of nationally reported cases, consistent with near-total surveillance collapse. After hostilities ended, a marked pediatric shift emerged, with the median age of infection in Tigray declining from 24.0 years during the conflict to 5.0 years in the post-conflict period (p < 0.0001), a pattern not observed in other regions. Compared with the pre-conflict baseline, the odds that a confirmed measles case was zero-dose were substantially higher during the conflict peak (aOR: 71.43; 95% CI: 14.1 - 1000.0) and remained elevated during post-conflict recovery (aOR: 2.49; 95% CI: 1.17 - 5.52). During the conflict peak, 50% of confirmed cases in Tigray lacked sex-disaggregated data.

**Conclusion:** The conflict in Tigray severely disrupted immunization services and surveillance, delaying detection of a large susceptible pediatric cohort. These findings underscore the need for age-targeted catch-up vaccination and resilient surveillance systems during post-conflict recovery.

**Key questions:** *What is already known?:*

- Armed conflict disrupts routine immunization and increases the risk of measles outbreaks in low- and middle-income countries.
- Conflict also weakens disease surveillance systems, which can delay outbreak detection and obscure the true burden of vaccine-preventable diseases.
- Measles is highly sensitive to disruptions in vaccination coverage and is often among the first diseases to resurge following health system breakdown.

*What are the new findings?:*

- During the 2020–2022 conflict in Tigray, measles surveillance reporting declined to near zero, despite rising national measles incidence.
- After the cessation of hostilities, measles cases in Tigray shifted sharply toward young children, revealing a large cohort of unvaccinated children that had accumulated during the conflict period.
- Compared with the pre-conflict period, confirmed measles cases in the post-conflict period had substantially higher odds of being zero-dose, indicating prolonged interruption of routine immunization services.

*What do the new findings imply?:*

- Surveillance data from conflict-affected settings may substantially underestimate disease burden during periods of active conflict, leading to delayed recognition of outbreaks.
- Post-conflict recovery strategies should prioritize age-stratified catch-up vaccination campaigns targeting children born during conflict periods, rather than relying solely on routine immunization services.
- Strengthening surveillance resilience in fragile and conflict-affected settings is essential to prevent delayed detection of measles and other vaccine-preventable disease outbreaks.

## 1. Introduction

Armed conflict remains one of the most persistent challenges to population health, not only through direct violence but through its indirect effects on health systems and essential services. While armed violence causes immediate morbidity and mortality, the longer-term burden is often driven by the disruption of healthcare delivery, supply chains, and public health infrastructure, resulting in excess preventable deaths well beyond the period of active hostilities[1,2]. Conflict-related displacement, overcrowding, and destruction of facilities further exacerbate vulnerability to infectious diseases, particularly among children[3,4]. Routine childhood immunization is especially susceptible to disruption, as it depends on stable logistics, functioning cold-chain systems, and consistent access to health workers - conditions that are frequently compromised during conflict[4,5].

Measles is among the most sensitive indicators of such systemic breakdowns. Due to its high transmissibility, sustained interruption of transmission requires very high population immunity, typically exceeding 95% vaccination coverage[6,7]. Even short-lived declines in immunization coverage can therefore result in outbreaks, making measles a reliable marker of gaps in routine service delivery[1,3]. In conflict-affected settings, disruptions to cold-chain infrastructure caused by power shortages or fuel constraints may render available vaccines ineffective, while displacement into crowded and resource-limited environments facilitates rapid transmission among undernourished and unvaccinated children[4,5]. As a result, measles outbreaks often reflect broader failures in health system functioning rather than isolated programmatic weaknesses.

Beyond immunization, armed conflict substantially weakens disease surveillance and outbreak response capacity. Effective surveillance depends on continuous reporting from peripheral health facilities to central authorities, supported by functional communication networks and laboratory systems[8 - 10]. In conflict zones, health facilities may be destroyed or abandoned, health workers displaced, and telecommunications disrupted, leading to prolonged interruptions in case reporting [10,11]. This produces periods of apparent epidemiological silence, during which reported incidence declines despite ongoing transmission[8,12]. Such surveillance gaps hinder timely response and obscure the true scale and distribution of disease, allowing susceptibility to accumulate undetected until services are partially restored[9,13].

Ethiopia has historically made substantial investments in strengthening its primary healthcare system, including expansion of the health extension program and improvements in routine immunization coverage[10,12]. Prior to 2020, the Tigray region was considered one of the better-performing regions in terms of health service delivery and surveillance capacity[11,12]. The outbreak of armed conflict in late 2020, however, precipitated a severe and localized collapse of health services. The conflict was characterized by widespread destruction of facilities, interruption of medical supply chains, and prolonged blackouts of electricity and telecommunications, effectively isolating the region from national health systems[12,14].

Following the cessation of large-scale hostilities in late 2022, attention has increasingly turned to the longer-term public health consequences of the conflict. While national disease control programs continued to operate in much of Ethiopia, Tigray experienced a prolonged period of limited-service restoration. As surveillance systems gradually resumed, emerging data suggested substantial changes in measles epidemiology that could not be readily explained by national trends alone. However, the extent to which these patterns reflect true shifts in disease burden versus delayed detection of previously hidden susceptibility remains poorly understood.

To address this gap, we analyzed seven years (2018 - 2024) of national measles case-based surveillance data to examine changes in measles epidemiology in Tigray before, during, and after the conflict. By comparing age distribution, surveillance completeness, and vaccination status in Tigray with the rest of Ethiopia, this study aims to quantify how conflict-related disruption of immunization and surveillance systems altered the observable epidemiology of measles and left a cohort of children vulnerable to post-conflict outbreaks.

## 2. Methodology

### 2.1. Study Design and Data Source

We conducted a retrospective, population-based cohort analysis utilizing the national measles case-based surveillance database of Ethiopia. This dataset is maintained by the Ethiopian Public Health Institute (EPHI) and serves as the primary repository for the Integrated Disease Surveillance and Response (IDSR) system. Surveillance data capture reported cases and do not estimate true incidence. The study period, spanning from January 2018 to December 2024, was selected to capture a robust pre-conflict baseline, the active conflict phase, and the subsequent recovery period.

### 2.2. Case Classification and Inclusion Criteria

Cases were systematically included based on the World Health Organization (WHO) and EPHI standardized case definitions [15,16]. Laboratory-confirmed cases were defined by clinical symptoms of measles (fever, maculopapular rash, and at least one of cough, coryza, or conjunctivitis) accompanied by a positive measles-specific IgM antibody test. Epidemiologically linked cases were those meeting the clinical definition with a direct temporal and geographical link, within 30 days, to a laboratory-confirmed case. To maintain the highest specificity for our demographic analysis and mitigate misclassification with other febrile rash illnesses during conflict-driven surveillance disruptions, we excluded cases classified as “Discarded” (IgM negative) or those identified solely as “Clinically Compatible” without laboratory or epidemiological linkage.

### 2.3. Defining Conflict Phases: The Temporal Framework

A central component of our analysis was the temporal stratification of the data into three “Conflict Phases,” allowing us to evaluate the conflict as a fundamental shift in the state of the health system. The Pre-Conflict period (2018 - 2019) represents a baseline of stable routine immunization and active surveillance. The Conflict Peak (2020 - 2022) is characterized by the onset of hostilities, widespread disruption of cold-chain infrastructure, and the systemic collapse of health reporting in affected areas [17,18]. Finally, the Post-Conflict/Recovery phase (2023 - 2024) marks the cessation of hostilities and the re-establishment of the national reporting system alongside supplemental immunization activities (SIAs).

### 2.4. Spatial Harmonization and Administrative Rescaling

During the study period, Ethiopia underwent significant administrative restructuring, notably the dissolution of the Southern Nations, Nationalities, and Peoples’ Region (SNNPR) into several smaller regional states. To ensure longitudinal consistency, we performed manual spatial harmonization by redistributing cases originally coded under SNNPR into the new regional entities - South Ethiopia, Central Ethiopia, South West Ethiopia, and Sidama - by cross-referencing zonal and woreda-level geographic identifiers with the 2023 administrative decree. This spatial harmonization was critical to prevent geographical artifacts from confounding regional comparisons and ensuring that national benchmarks remained accurate over time.

### 2.5. Demographic disaggregation and vaccination status classification

To characterize demographic shifts within the affected population, we summarized patient characteristics using descriptive statistics stratified by vaccination status, categorizing cases into Zero-Dose (no doses prior to infection) as a proxy for the “unreached” population, and ≥1 Dose as an operational proxy for prior contact with immunization services that did not result in effective protection at the population level, plausibly reflecting conflict-related disruptions such as delayed schedules, cold-chain instability, documentation gaps, or waning immunity rather than direct vaccine failure [11]. These cohorts were disaggregated by biological sex-standardized to title-case to address reporting “sparsity” and three age brackets (infants <9 months, target pediatric 9–59 months, and older cohorts 5+ years), with continuous age analysis utilizing the median and interquartile range (IQR) to account for the non-normal, multi-modal distribution observed in the raw data. Notably, missing demographic data, which exceeded 50% for biological sex in Tigray during the conflict peak, was treated as a sentinel indicator of surveillance reporting strain rather than simple random omission, and was addressed through a combination of complete-case analysis for disaggregation and stratified median imputation for age to mitigate sentinel bias.

### 2.6. Addressing Informative Missingness and Sentinel Bias

Surveillance data in conflict zones frequently suffer from informative missingness, where the probability of data being missing is directly related to conflict intensity. During the conflict peak, surveillance in Tigray was largely restricted to mobile adults or individuals able to access the few functioning urban health facilities, resulting in sentinel bias. To address this, we employed a stratified median imputation strategy. Missing age values were replaced using the median age calculated within each region and conflict phase, a conservative approach chosen to preserve distributional robustness in the presence of right-skewed age distributions and to minimize distortion from adult-dominated reporting during the conflict period. This approach reduced the likelihood that conflict-related reporting patterns would bias age distributions observed during the recovery phase.

All ages were converted to decimal years, incorporating months for infants, to improve precision in the 0 - 5-year age group, where the epidemiological impact of missed routine immunization is most pronounced.

### 2.7. Statistical Analysis and Interaction Modeling

To mathematically evaluate the shift in transmission, we employed a Two-Way Analysis of Variance (ANOVA) framework within a linear interaction model (Age ∼ Region x Phase). This model assessed the significance of the interaction between the region and the conflict phase to determine if Tigray’s demographic shift was a localized anomaly compared to the national baseline. Non-parametric analysis was further supported by the Wilcoxon Rank Sum test to compare median ages between the Conflict Peak and Post-Conflict periods[19].

Finally, to quantify the odds that a confirmed measles case was zero-dose, we developed a multivariable logistic regression model to calculate adjusted Odds Ratios (aOR) and 95% Confidence Intervals (CI) for Zero-Dose (unvaccinated) status. The model was adjusted for the primary exposure (Conflict Phase) and potential confounders, including age and sex. By setting the Pre-Conflict phase as the reference group, we isolated the relative increase in the likelihood of a confirmed case being unvaccinated during the post-conflict resurgence. These estimates are conditional on reported cases and do not represent population-level vaccination coverage. All analyses were conducted in R (Version 4.5.1) using the tidyverse, lubridate, and broom packages[20]. This study is reported in accordance with the STROBE reporting guidelines for observational studies.

## 3. Results

### 3.1. The Reporting void and surveillance blackout

The analysis reveals a profound divergence between national epidemiological trends and regional reporting in Tigray during the conflict period. Between 2018 and 2020, Tigray maintained a stable reporting baseline, contributing up to 4.5% of the national measles case-load. However, following the onset of hostilities, a “statistical blackout” occurred: while national cases surged from 1,884 in 2021 to 8,482 in 2022, Tigray’s reported cases plummeted to a low of just 1 case in 2022 (Table 1, Figure 1).

**Table 1.** Characteristics of Measles Cases by Region and Conflict Period.

| Year | National cases | Tigray cases<br>n (%) | Status |
| --- | --- | --- | --- |
| 2018 | 1,489 | 22 (1.48) | Baseline |
| 2019 | 2,628 | 55 (2.09) | Baseline |
| 2020 | 1,865 | 84 (4.5) | Conflict Onset |
| 2021 | 1,884 | 2 (0.11) | System Collapse |
| 2022 | 8,482 | 1 (0.01) | System Collapse |
| 2023 | 23,762 | 57 (0.24) | Resurgence/Recovery |
| 2024 | 29,756 | 24 | Resurgence/Recovery |

**Table 2.**
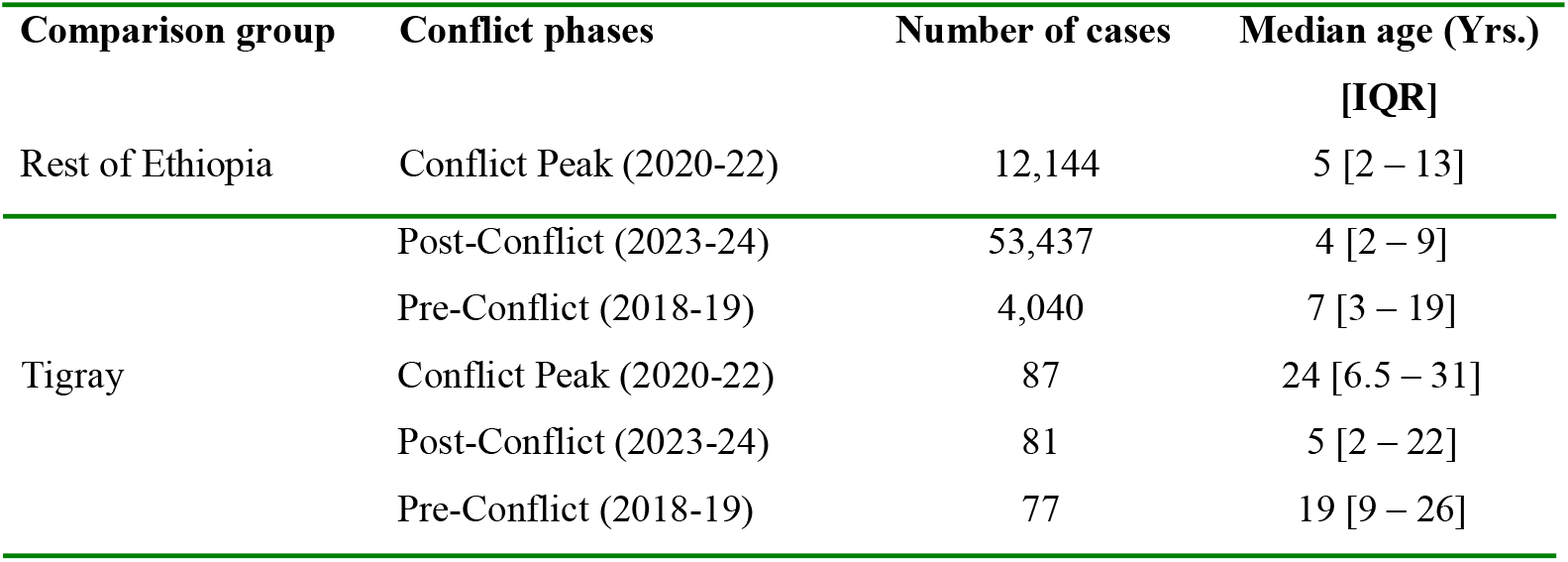
Age structure of measles cases (Medians and IQR) by conflict phases.

| Comparison group | Conflict phases | Number of cases | Median age (Yrs.)<br>[IQR] |
| --- | --- | --- | --- |
| Rest of Ethiopia | Conflict Peak (2020-22) | 12,144 | 5 [2 – 13] |
| Tigray | Post-Conflict (2023-24) | 53,437 | 4 [2 – 9] |
|  | Pre-Conflict (2018-19) | 4,040 | 7 [3 – 19] |
|  | Conflict Peak (2020-22) | 87 | 24 [6.5 – 31] |
|  | Post-Conflict (2023-24) | 81 | 5 [2 – 22] |
|  | Pre-Conflict (2018-19) | 77 | 19 [9 – 26] |

**Figure 1.**
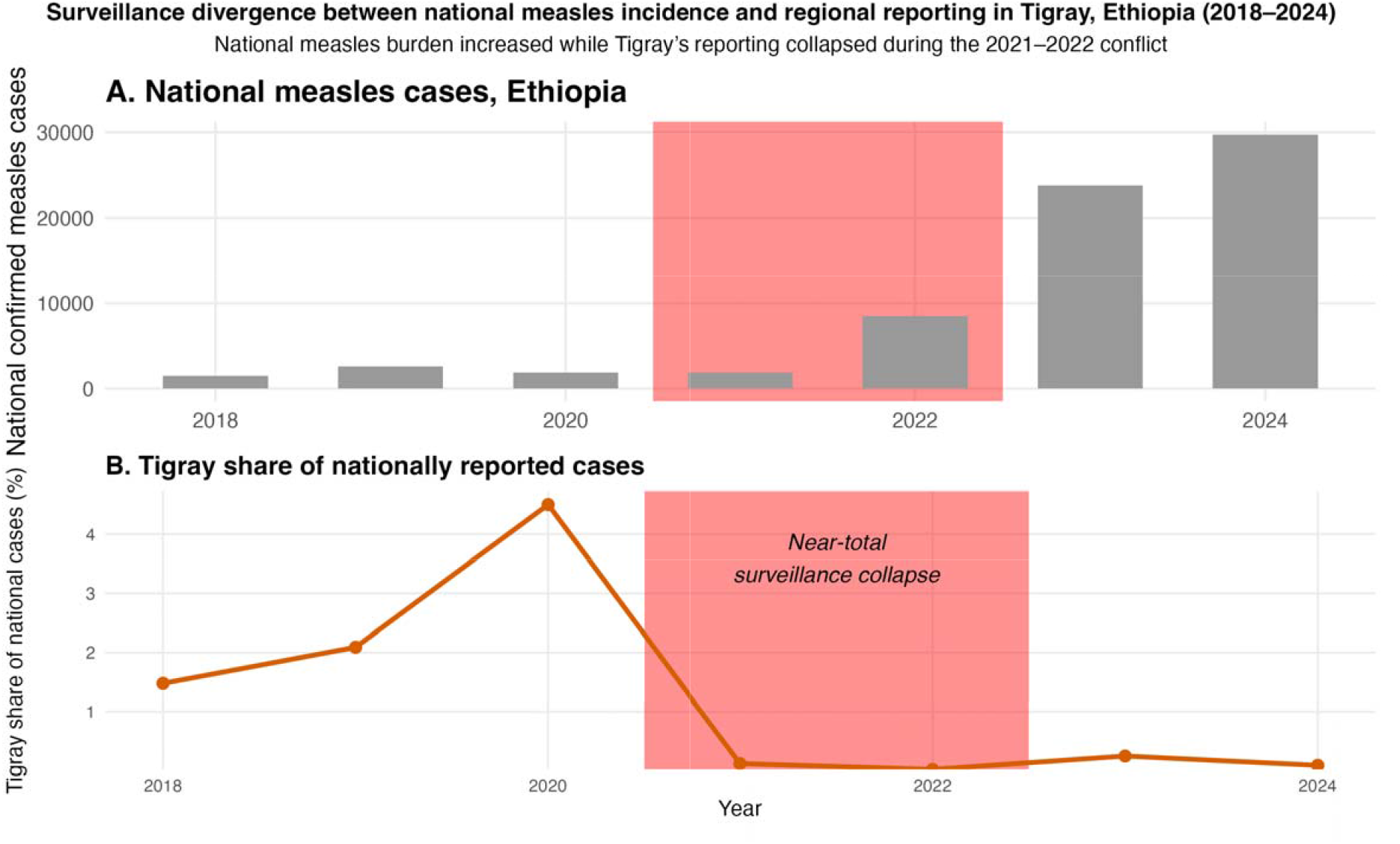
The measles surveillance divergence between Ethiopia and Tigray region (national surge vs. regional blackout)

This represents a drop in regional reporting efficiency to 0.01%, signaling a near-total collapse of the IDSR system during the conflict peak. Despite a nationwide epidemic expansion in 2023 and 2024 (reaching nearly 30,000 cases), Tigray’s reporting share remained suppressed at <0.1%, indicating a lingering surveillance gap during the recovery phase. Importantly, these reductions in reported cases should be interpreted as surveillance failure rather than evidence of reduced measles transmission during the conflict period. (Figure 1, Table 1).

### 3.2. The Demographic shift in age distribution

Statistical analysis of the age distribution confirms a substantial demographic shift in the outbreak profile across the three defined phases. During the peak of the conflict (2020 - 2022), the median age of laboratory-confirmed measles cases in Tigray rose to 24.0 years, with a wide interquartile range (IQR: 16.0–32.0 years). This unusually adult-centered and diffuse distribution is unlikely to reflect a true change in measles epidemiology, but rather a profound distortion in case ascertainment, whereby the surveillance system predominantly captured mobile, symptomatic adults or cases presenting to the few functioning urban health facilities. In contrast, pediatric transmission - historically the core of measles epidemiology - was largely absent from official records during this period.

Following the partial and gradual restoration of health services after the cessation of hostilities (2023 - 2024), the median age of infection dropped sharply to 5.0 years, and the age distribution became highly compressed within early childhood (IQR: 2.0 - 9.0 years), revealing a classic pediatric outbreak pattern. This abrupt reversion is unlikely to reflect a true epidemiologic transition. Instead, it indicates delayed detection of accumulated susceptibility. The contrast between the conflict and post-conflict age distributions was highly significant (Wilcoxon rank-sum test, *p* < 0.0001), reinforcing the magnitude of this demographic reversal.

Comparison with the neighboring Amhara region - where a consistently pediatric-focused age distribution was maintained throughout the study period - further highlights the uniqueness of the Tigray experience. The absence of a similar demographic distortion in Amhara supports the interpretation that the observed shift in Tigray was driven by conflict-related disruptions to immunization and surveillance rather than broader national or seasonal changes in measles transmission. Together, these findings illustrate how prolonged service collapse can mask pediatric outbreaks and allow susceptibility to accumulate, only to be revealed once minimal surveillance and healthcare access are restored (Figure 2).

**Figure 2.**
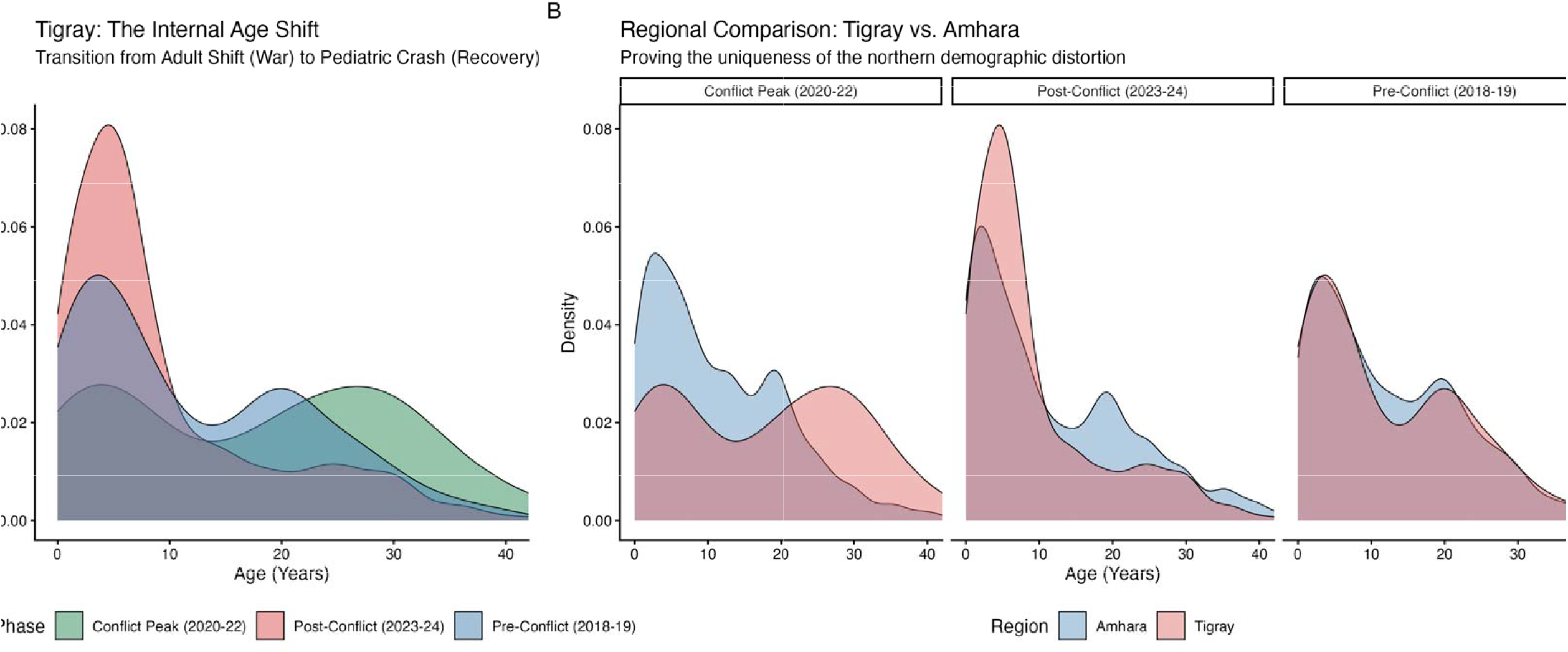
Age distribution of confirmed measles cases during conflict and post-conflict recovery in Tigray, Ethiopia. (A) Kernel density plots showing the age distribution of laboratory-confirmed measles cases in Tigray from 2018 to 2024. The distribution shifts from a predominantly pediatric pattern before the conflict to a broader, adult-skewed profile during the conflict period (2020–2022), followed by a marked return to younger age groups in the post-conflict period (2023–2024). (B) Comparative age distributions between Tigray and the neighboring Amhara region across the same periods. While Amhara demonstrates a stable, pediatric-dominated age profile throughout, Tigray shows a pronounced temporal distortion during the conflict, consistent with disruption of routine immunization and health service access.

To determine if the demographic shifts observed in Tigray were merely a reflection of national trends or a localized consequence of the conflict, we performed a Two-Way Analysis of Variance (ANOVA) examining the interaction between Region and Conflict phases. The model demonstrated a highly significant interaction effect (*F* (2, 69,860) = 94.3, *p* < 0.0001), providing definitive evidence that the age of infection evolved differently in Tigray than in the rest of Ethiopia.

The linear interaction model further clarified these differences through the estimated coefficients. The interaction term for Tigray during the post-conflict period (2023 - 2024) was strongly and independently associated with a reduction in age at infection (β = -8.22, *p* < 0.0001). This effect indicates that the pronounced decline in the age of reported cases in Tigray - returning to a predominantly pediatric profile - was a statistically distinct phenomenon. The magnitude and direction of this interaction support the interpretation that the adult-skewed age distribution observed during the conflict represented a localized distortion in case detection rather than a true shift in transmission dynamics. Importantly, this correction occurred only after partial restoration of health services and surveillance and was not observed in other regions, where a stable pediatric age profile was maintained throughout the study period (table 3).

**Table 3.** ANOVA table showing the interaction between Region and Conflict phases.

| Source of Variation | Sum of Squares | Degrees of freedom | Mean Squares | F-value | Pr. (>F) |
| --- | --- | --- | --- | --- | --- |
| Region (Tigray vs. Rest) | 22,056 | 1 | 22,056 | 306.8 | $< 2.2 \times 10^{-16}$ *** |
| Phase (Conflict Phases) | 67,812 | 2 | 33,906 | 471.6 | $< 2.2 \times 10^{-16}$ *** |
| Region × Phase Interaction | 13,564 | 2 | 6,782 | 94.3 | $< 2.2 \times 10^{-16}$ *** |
| Residuals | 5,022,934 | 69,860 | 71.9 |  |  |

\*\*\* $P < 0.0001$

### 3.3. Comparative Analysis of Zero-Dose Status

The proportion of Zero-Dose cases - defined as children who had received no doses of measles-containing vaccine prior to infection - provides a direct measure of the health system’s inability to provide basic primary care. In Tigray, the Zero-Dose proportion among reported cases was historically very low at 10.2% (Pre-Conflict), indicating a highly inclusive and effective immunization reach. During the Conflict Peak, this tripled to 46.4%, a clear reflection of the total cessation of services during the hostilities.

The most revealing finding, however, occurs in the post-conflict phase. In Tigray, the Zero-Dose proportion among cases fell to 13.2%, nearly returning to its pre-war baseline. In contrast, the Rest of Ethiopia saw its Zero-Dose proportion rise steadily from 23.7% to 36.6% over the same period. This suggests two very different public health crises. In the Rest of Ethiopia, the problem is a growing population of children who have never been reached. In Tigray, the system is successfully reaching children (only 13.2% are Zero-Dose), yet measles transmission remains high. This discrepancy suggests that post-conflict measles transmission in Tigray may reflect residual operational weaknesses in immunization delivery such as delayed schedules, incomplete dosing, or compromised service continuity rather than failures of population reach alone (table 4).

**Table 4.** Comparison of zero dose vaccination proportion between Tigray and the rest of Ethiopia by conflict phases.

| Comparison Group | Conflict Phase | Zero-Dose (%) | 95% Confidence Interval |
| --- | --- | --- | --- |
| **Tigray** | Pre-Conflict | 10.2% | 6.9 - 14.9 |
|  | Conflict Peak | 46.4% | 37.9 - 55.1 |
|  | Post-Conflict | 13.2% | 10.2 - 17.0 |
| **Rest of Ethiopia** | Pre-Conflict | 23.7% | 22.8 - 24.6 |
|  | Conflict Peak | 28.4% | 27.8 - 29.0 |
|  | Post-Conflict | 36.6% | 36.2 - 37.0 |

To isolate the impact of the conflict on immunization coverage while controlling for demographic confounders, we employed a multivariable logistic regression model with the Pre-Conflict period (2018 - 2019) as the reference baseline. The analysis revealed a profound temporal escalation in the risk of zero-dose status. During the conflict peak (2020 - 2022), the odds of a confirmed measles case being Zero-Dose were 71.4 times higher than the pre-war baseline (aOR: 71.43; 95% CI: 14.1 - 1000.0; p<0.001), highlighting a near-total cessation of routine immunization services. Although recovery efforts were initiated in the post-conflict phase (2023 - 2024), the risk of being Zero-Dose remained significantly elevated at 2.5 times the pre-conflict level (aOR: 2.49; 95% CI: 1.17–5.52; *p*=0.020), underscoring the lingering immunity gap within the region.

Age functioned as a significant continuous predictor of vaccination status (aOR: 0.95; p<0.001), where each additional year of age was associated with a 5% decrease in the likelihood of being Zero-Dose. This trend likely reflects the cumulative opportunities for vaccination afforded to older cohorts prior to the healthcare system’s collapse. Conversely, biological sex was not a significant predictor of immunization status (*p*=0.474), with no statistically significant difference observed between male and female cases. This suggests that the susceptibility gap in Tigray was driven by systemic geographic and temporal barriers such as the regional blockade and cold-chain destruction rather than gender-based disparities in healthcare seeking (Table 5).

**Table 5.**
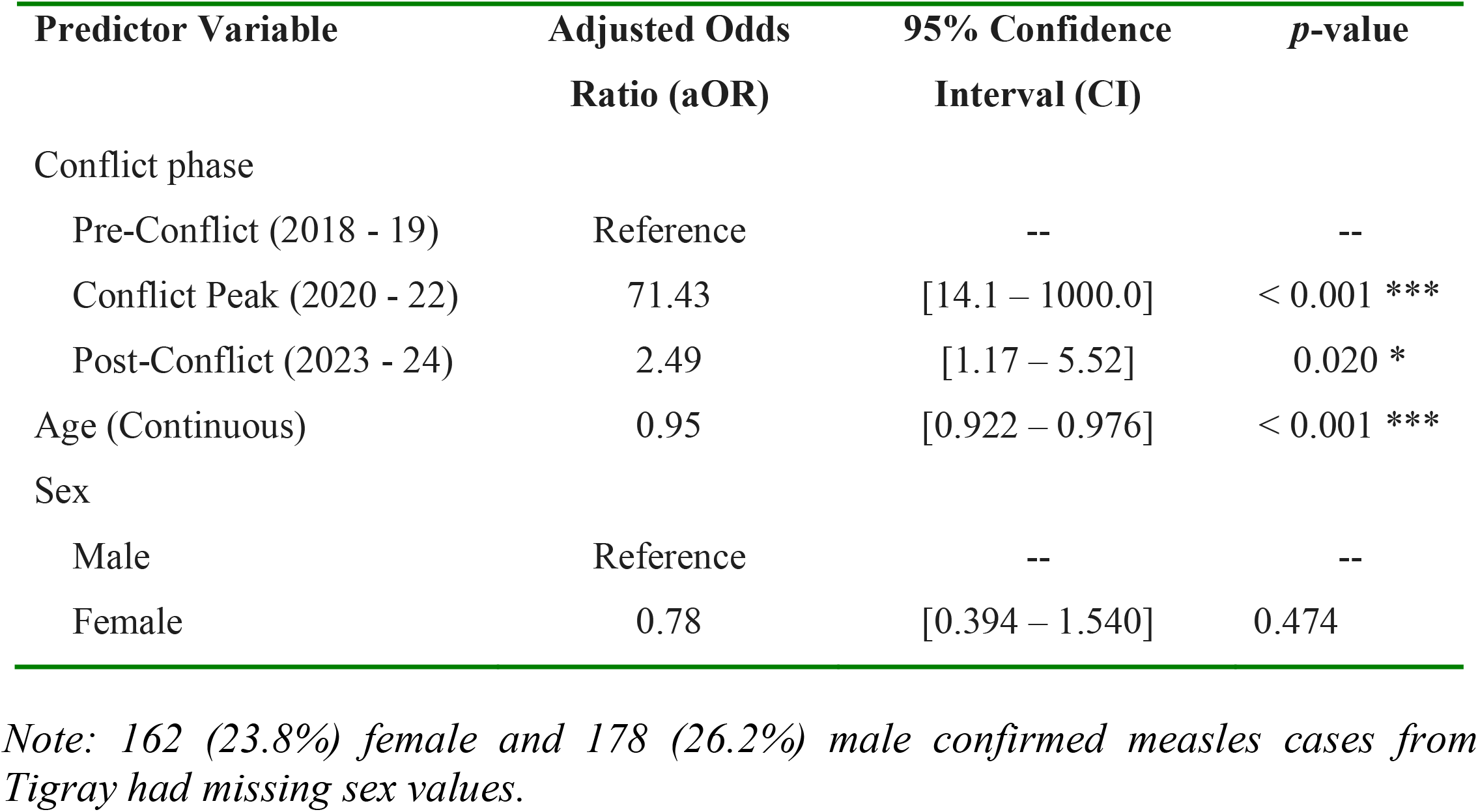
Multivariable logistic regression analysis of factors associated with Zero-Dose (Unvaccinated) status among confirmed measles cases in Tigray region.

## 4. Discussion

The demographic shift observed in Tigray represents a clear epidemiological signature of severe health system disruption. During the conflict period, the shift toward an adult-dominated median age does not reflect a biological change in measles transmission but is more plausibly explained by a surveillance filtering effect resulting from the collapse of pediatric healthcare access and routine reporting mechanisms [21,22]. Under these conditions, the IDSR system primarily captured mobile, symptomatic adults who were able to reach the limited functioning urban health facilities, while pediatric cases in rural and inaccessible areas remained largely undetected. The subsequent decline in the median age of infection to five years following the cessation of hostilities reflects the delayed detection - or “unmasking” - of accumulated susceptibility among children who missed routine immunization during the conflict period[23,24]. Similar patterns of concealed pediatric transmission have been documented in other conflict-affected settings, including Syria, where reporting efficiency temporarily decoupled from true disease burden during periods of insecurity [25].

The markedly increased odds of zero-dose status among confirmed cases in the post-conflict period further quantify the extent of this susceptibility gap. While these estimates are conditional on reported cases and should not be interpreted as population-level coverage measures, the magnitude of the observed disparity indicates a substantial interruption of routine immunization services. In other humanitarian crises, including Yemen and the Democratic Republic of the Congo, conflict-related disruptions have been associated with two- to five-fold increases in zero-dose prevalence [17,26,27]. The considerably larger effect observed in Tigray suggests a near-complete breakdown of vaccine delivery and likely cold-chain functionality during the conflict period, resulting in a prolonged accumulation of susceptible cohorts [4,28]. In this context, measles case detection patterns reflect the spatial and temporal distribution of health service access rather than solely underlying transmission dynamics, consistent with prior evidence from settings experiencing prolonged blockades or service collapse [29].

The scale of the observed susceptibility gap indicates that restoration of routine immunization services alone is unlikely to be sufficient to prevent future outbreaks. Instead, targeted, age-stratified Supplemental Immunization Activities (SIAs) are required to address immunity gaps among children born during the conflict period, particularly those born between 2020 and 2023 who were systematically missed by routine services[30,31]. Strengthening the resilience of the surveillance system is equally critical. Decentralized surveillance approaches, including community-based reporting and diagnostic capacity that does not rely exclusively on centralized laboratory infrastructure, may help reduce future reporting voids in fragile settings [21,32]. Integration of vaccination and surveillance efforts with existing social protection mechanisms, such as the Productive Safety Net Program (PSNP), may further improve access to marginalized and hard-to-reach populations [31].

A key strength of this study is the use of a national comparative framework, allowing the epidemiological patterns observed in Tigray to be distinguished from broader national trends. By comparing Tigray with the rest of Ethiopia, we were able to isolate the conflict as the primary driver of the observed age redistribution, rather than attributing these changes to national shifts in measles epidemiology or seasonality [22]. Nevertheless, several limitations must be acknowledged. The absence of surveillance during 2021 - 2022 introduces substantial selection bias, with reported cases likely representing a small and atypical subset of the true burden, skewed toward mobile and urban populations. Although stratified median imputation was used to address missing age data, this approach cannot fully reconstruct epidemiological patterns lost during periods of complete reporting disruption [33]. In addition, the absence of reliable case-fatality data limits assessment of the mortality impact associated with the post-conflict pediatric resurgence.

## 5. Conclusion

The 2020 - 2022 conflict in Tigray dismantled years of immunization gains, leading to a profound increase in zero-dose status and a conflict-specific shift in the age distribution of measles cases that revealed a large, previously undetected pediatric epidemic. Measles proved to be a sensitive indicator of health system breakdown: when surveillance collapsed, transmission persisted silently among vulnerable populations. Addressing this accumulated susceptibility will require urgent implementation of multi-cohort catch-up vaccination strategies and the development of decentralized, conflict-resilient surveillance systems to prevent future high-mortality outbreaks during post-conflict recovery.

## Abbreviations

ANOVA: Analysis of Variance
EPHI: Ethiopian Public Health Institute
CI: Confidence Interval
IQR: Interquartile Range
IDSR: Integrated Disease Surveillance and Response
PSNP: Productive Safety Net Program
SIA: Supplemental Immunization Activity
WHO: World Health Organization

## Acknowledgements

We would like to extend our gratitude to the Ethiopian Public Health Institute for providing the data used in this study. We would also like to thank the colleagues in the Ethiopian Public Health Institute for providing the necessary support in addressing data quality and linking the measles data with geographic coordinates. We further acknowledge the financial support provided by Sungkyunkwan University, BK21 FOUR (Graduate School Innovation), and the Ministry of Education and Ministry of Science & ICT of the Republic of Korea. Finally, we would also like to thank all the coauthors who reviewed and provided critical comments to improve the manuscript.

## Author contributions

HMW, YB and SWL developed the study protocol, supervised the execution of the study, took part in data analysis, and wrote the first draft of the manuscript. HMW, YB and SWL conducted statistical analysis and modelling and generated the figures and tables for the manuscript. YB, and AR critically reviewed the manuscript. All the authors have also read and approved the final version of the manuscript.

## Data availability

The data used in this study are not publicly available due to national data protection regulations but can be obtained from the corresponding author upon reasonable request and with permission from the relevant national authorities.

## Ethical consideration

This study used secondary, anonymized measles surveillance data routinely collected by the Public Health Emergency Management (PHEM) unit of the Ethiopian Public Health Institute for emergency response and public health monitoring. All direct personal identifiers, including names and specific residential addresses, were removed prior to analysis to ensure confidentiality and data protection, in accordance with the principles of the Declaration of Helsinki.

As the study involved retrospective analysis of fully de-identified data, it was exempt from full institutional review board review. Authorization to access and analyze the surveillance data was obtained from the relevant national authorities. All results are reported in aggregated form to prevent the identification of individual cases.

## Patient and Public Involvement

Patients and members of the public were not directly involved in the design, conduct, reporting, or dissemination plans of this research. This study was based on the secondary analysis of routinely collected, anonymized national measles surveillance data used for public health monitoring and outbreak response. As such, no individual-level patient engagement was feasible or required.

## Consent for publication

Not applicable.

